# A model-driven machine learning approach for personalized kidney graft risk prediction

**DOI:** 10.1101/2023.10.01.23296293

**Authors:** Symeon V. Savvopoulos, Irina Scheffner, Andreas Reppas, Wilfried Gwinner, Haralampos Hatzikirou

## Abstract

Graft failure after renal transplantation is a multifactorial process. Predicting the risk of graft failure accurately is imperative since such knowledge allows for identifying patients at risk and treatment personalization. In this study, we were interested in predicting the temporal evolution of graft function (expressed as estimated glomerular filtration rate; eGFR) based on pretransplant data and early post-operative graft function. Toward this aim, we developed a tailored approach that combines a dynamic GFR mathematical model and machine learning while taking into account the corresponding parameter uncertainty. A cohort of 892 patients was used to train the algorithm and a cohort of 847 patients for validation. Our analysis indicates that an eGFR threshold exists that allows for classifying high-risk patients. Using minimal inputs, our approach predicted the graft outcome with an accuracy greater than 80% for the first and second years after kidney transplantation and risk predictions were robust over time.

## 1. Introduction

The challenge in the management of patients with chronic organ disease is to prevent progressive organ deterioration and ultimate failure. Most chronic diseases are characterized by a dynamic process of injury and repair responses inherent to the specific disease present. This natural course may be modulated by a complex, time-variant interplay of disease-modifying factors, co-morbidity conditions, and potential clinical interventions. Rational appreciation of this complex interplay is the key for an individualized patient treatment in terms of diagnostic measures and therapeutic decisions. This includes recognition and weighing of relevant causative and non-causative risk factors, their potential interaction, and estimation of putative therapeutic effects with respect to the clinical disease course and the mid- and long-term outcomes^1^.

Kidney transplantation is a prime paradigm of a complex chronic disease with a dynamic course. For patients with end-stage renal failure, kidney transplantation is the optimal choice of treatment as it restores renal function significantly. The degree of renal function is usually estimated by the glomerular filtration rate (GFR)^2^. In particular, the GFR is determined by the quality of the transplanted organ. After transplantation, various factors determine the further evolution of GFR, including peri- and post-operative complications and successful treatment of these complications. In an uneventful post-transplant course, kidney grafts can last several decades without relevant loss of GFR^3^. However, in many cases, repeated injuries lead to irreversible graft injury and progressive loss of GFR^4,5^. Injuries to the graft include acute and chronic T cell- and antibody-mediated rejection, drug toxicity, infections, recurrence of renal disease, and effects from patients’ co-morbidities which may have a cumulative impact on the transplantat outcome. Appropriate therapeutic intervention may successfully treat these complications but often leads only to partial or no improvement. Hence, despite all previous diagnostic and therapeutic progress, the mean kidney graft survival is limited to approximately 15 years^6,7^. Thus, besides timely recognition of the injuries to the graft, the challenge is to have a precise understanding of the nature and extent of heterogenous and complex injury factors on the outcome, in this case, the graft function (GFR).

Common computational approaches for the assessment of chronic disease risk factors and outcomes are regression analysis and Cox proportional hazard analysis, which assign a certain effect value to the putative risk factors and numerically estimate the outcome of interest in a given study population^8^. In such approaches, the patient’s status is captured at a specific point in time by choosing particular risk factors, and in turn, an estimate for the future course is predicted. By nature, these approaches cannot truly integrate time-variant, sequential events of dynamic disease processes into the model, besides from condensing these multiple events into suitable factor variables. Also, the appropriate estimation of potential interactions between different factors that modulate the GFR is challenging. Armero et al. developed a model for predicting the GFR in children with chronic kidney disease. A Bayesian analysis estimated the joint model’s parameters, hyperparameters, and random effects^9^. The drawback of the proposed model is its struggle to accurately predict disease progression for children with serious or irregular profiles due to the complex and variable nature of chronic kidney disease, as evidenced by its deficiency in identifying such cases and its inability to fully explain the observed variability in progression patterns. Limitations of this model included inaccurate prediction of disease progression in children with severe graft impairment or with irregular functionional changes and latent disease states, thus making identification of these cases virtually impossible despite application of advanced approaches like Markov Mixture Models.

Recently, a sequence-to-sequence deep learning algorithm was developed to predict the patient-specific expected reference range of the GFR by using serial GFR measurements from the first 3 months posttransplantation^10^ The algorithm was shown to be helpful identifying GFR deviations in the individual that indicated truly anomalous values and thus, need of potential further diagnostic work-up. Yet, a large number of patient’s GFR measurements was required for this assessment and the algorithm could not estimate the stability properties of this value, i.e. whether the GFR will change over time or stay invariant.

Using data from a large cohort of kidney transplant patients and employing machine learning algorithms^11^, we aimed to build a dynamic and expandable model of the graft function after kidney transplantation. In this paper, we developed a framework that assesses the risk of graft failure in the first and second year using a small number of pre-operative clinical data and two consecutive GFR measurements after transplantation.

## 2. Results

### 2.1. Donor’s age, post-operative GFR values and GFR speed provide a satisfactory estimate of the first-year graft function

Table 1 and Table 2 include the patients’ clinical information for the training and validation groups, respectively. We have tested the difference of our variables between different patient groups using the χ^2^ test for categorical variables and Mann-Whittney test for continuous variables with p-value ≤ 0.05. In the training group, patients with rejection were younger, had a lower GFR within the first six weeks after transplantation and had more often pre-formed antibodies at transplantation. In the validation cohort, patients with rejection were older, were more often retransplanted and received more often organs from older and deceased donors. Patients with one or multiple rejections received more often organs from a female donor, and patients with one rejection were more often male. Patients without rejection had a more favorable immunological risk profile, with fewer HLA mismatches and lower prevalence of preformed antibodies and blood transfusions before transplantation. Patients with rejection more often lacked immediate graft function after transplantation and had a lower GFR within the first six weeks.

**Table 1:** Baseline characteristics of study participants in the training group (N_1_). Parentheses show the proportions.

|  | No rejection | One rejection | Multiple rejections | p-values |
| --- | --- | --- | --- | --- |
| N | 362 | 187 | 343 |  |
| Recipient age (yrs) | 51.21 ± 12.94 | 50.24 ± 13.21 | 48.00 ± 13.21 | **0.00** |
| Recipient sex |  |  |  |  |
| Male | 212 (59) | 115 (61) | 202 (59) | 0.17 |
| Female | 150 (41) | 72 (39) | 141 (41) | 0.19 |
| Donor age (yrs) | 47.82 ± 15.69 | 49.02 ± 16.29 | 49.18 ± 15.07 | 0.48 |
| Donor's sex |  |  |  |  |
| Male | 192 (53) | 99 (53) | 170 (50) | 0.18 |
| Female | 170 (47) | 88 (47) | 173 (50) | 0.17 |
| Origin of the renal graft |  |  |  |  |
| Deceased | 305 (84) | 162 (87) | 291 (85) | 0.07 |
| Living | 57 (16) | 25 (13) | 52 (15) | 0.48 |
| Cold Ischemia Time (hrs) | 14.14 ± 7.18 | 14.79 ± 7.32 | 14.79 ± 7.68 | 0.66 |
| Initial function of the graft |  |  |  |  |
| No | 82 (23) | 50 (27) | 127 (37) | 0.15 |
| Yes | 280 (77) | 137 (73) | 216 (63) | 0.07 |
| HLA-mismatch at locus A |  |  |  |  |
| 0 | 171 (47) | 80 (43) | 149 (43) | 0.15 |
| 1 | 142 (39) | 72 (38) | 145 (42) | 0.21 |
| 2 | 49 (14) | 35 (19) | 49 (14) | 0.85 |
| HLA-mismatch at locus B |  |  |  |  |
| 0 | 124 (34) | 56 (30) | 104 (30) | 0.24 |
| 1 | 158 (44) | 93 (50) | 157 (46) | 0.32 |
| 2 | 80 (22) | 38 (20) | 82 (24) | 0.36 |
| HLA-mismatch at locus DR |  |  |  |  |
| 0 | 165 (46) | 67 (36) | 103 (30) | 0.09 |
| 1 | 147 (41) | 91 (49) | 178 (52) | 0.21 |
| 2 | 50 (14) | 29 (15) | 62 (18) | 0.52 |
| Level of pre-formed antibodies (%) |  |  |  |  |
| >0% | 23 (6) | 11 (6) | 30 (9) | 0.62 |
| 0% | 339 (94) | 176 (94) | 313 (91) | **0.05** |
| Blood transfusions before transplantation |  |  |  |  |
| unknown | 54 (15) | 20 (11) | 60 (17) | 0.31 |
| yes | 107 (30) | 49 (26) | 99 (29) | 0.27 |
| no | 201 (55) | 118 (63) | 184 (54) | 0.28 |
| Number of pregnancies |  |  |  |  |
| no pregnancies | 248 (69) | 142 (76) | 241 (70) | 0.16 |
| yes | 101 (28) | 41 (22) | 88 (26) | 0.24 |
| unknown | 13 (4) | 4 (2) | 14 (4) | 0.72 |
| Previous transplantations |  |  |  |  |
| 1-2 | 45 (12) | 17 (9) | 55 (16) | 0.33 |
| none | 317 (88) | 170 (91) | 288 (84) | 0.07 |
| Highest GFR within 6 weeks after transplantation | 61.13 ± 24.08 | 62.00 ± 23.16 | 55.89 ± 24.68 | **0.00** |

**Table 2:** Baseline characteristics of study participants in the validation group (N_2_). Parentheses show the proportions.

|  | No rejection | One rejection | Multiple rejections | p-values |
| --- | --- | --- | --- | --- |
| N | 426 | 159 | 262 |  |
| Recipient age (yrs) | 49.75 ± 14.08 | 53.14 ± 12.86 | 50.69 ± 13.73 | **0.045** |
| Recipient sex |  |  |  |  |
| Male | 267 (63) | 105 (66) | 161 (61) | **0.01** |
| Female | 159 (37) | 54 (34) | 101 (39) | **0.05** |
| Donor age (yrs) | 51.19 ± 14.74 | 52.56 ± 15.76 | 53.73 ± 15.32 | 0.10 |
| Donor's sex |  |  |  |  |
| Male | 200 (47) | 65 (41) | 105 (40) | **0.01** |
| Female | 226 (53) | 94 (59) | 157 (60) | **0.05** |
| Origin of the renal graft |  |  |  |  |
| Deceased | 201 (47) | 125 (79) | 185 (71) | 0.35 |
| Living | 225 (53) | 34 (21) | 77 (29) | **0.00** |
| Cold Ischemia Time (hrs) | 9.11 ± 6.06 | 9.72 ± 5.09 | 9.09 ± 5.49 | 0.22 |
| Initial function of the graft |  |  |  |  |
| Yes | 378 (89) | 139 (87) | 211 (81) | **0.00** |
| no | 48 (11) | 20 (13) | 51 (19) | 0.44 |
| HLA-mismatch at locus A |  |  |  |  |
| 0 | 172 (40) | 52 (33) | 69 (26) | **0.01** |
| 1 | 182 (43) | 76 (48) | 132 (50) | 0.09 |
| 2 | 72 (17) | 31 (19) | 61 (24) | 0.40 |
| HLA-mismatch at locus B |  |  |  |  |
| 0 | 111 (26) | 34 (21) | 45 (17) | **0.05** |
| 1 | 196 (46) | 78 (49) | 115 (44) | **0.04** |
| 2 | 119 (28) | 47 (30) | 102 (39) | 0.17 |
| HLA-mismatch at locus DR |  |  |  |  |
| 0 | 165 (39) | 54 (34) | 66 (25) | **0.01** |
| 1 | 195 (46) | 71 (45) | 132 (50) | **0.04** |
| 2 | 66 (15) | 34 (21) | 64 (25) | 0.52 |
| Level of pre-formed antibodies (%) |  |  |  |  |
| >0% | 61 (14) | 22 (14) | 41 (16) | 0.36 |
| 0% | 365 (86) | 137 (86) | 221 (84) | **0.00** |
| Blood transfusions before transplantation |  |  |  |  |
| unknown | 68 (16) | 23 (14) | 60 (23) | 0.28 |
| yes | 82 (19) | 36 (23) | 52 (20) | 0.34 |
| no | 276 (65) | 100 (63) | 150 (57) | **0.01** |
| Number of pregnancies |  |  |  |  |
| no pregnancies | 313 (73) | 114 (72) | 183 (70) | **0.00** |
| yes | 85 (20) | 38 (24) | 58 (22) | 0.36 |
| unknown | 28 (7) | 7 (4) | 21 (8) | 0.51 |
| Previous transplantations |  |  |  |  |
| 1-2 | 41 (10) | 18 (11) | 36 (14) | 0.60 |
| none | 385 (90) | 141 (89) | 226 (86) | **0.00** |
| Highest GFR within 6 weeks after transplantation | 66.24 ± 25.50 | 63.29 ± 23.25 | 58.79 ± 23.63 | **0.00** |

First, we attempted to predict the GFR at 365 days post-transplantion in the training cohort. We used a linear regression analysis, which allowed not only to predict but also to identify the important determinant variables. Our results show that the highest GFR within the first 6 weeks and the GFR speed along with the donor’s age are sufficient to predict the GFR at 365 days. The R^2^ values of the multi-linear regression results were 0.68 for the non rejection patients, 0.57 for patients with one rejection, and 0.49 for patients with multiple rejections. Details regarding the regression approach are included in Section 2 of the SI (Section 2 of the SI including Figures S.1.-S.4. and Tables S.1. and S.2.).

### 2.2. A GFR threshold exists with respect to graft survival

Our GFR predictive model invoked the existence of a critical GFR threshold that is related to graft loss. To our knowledge, in the current literature the existence of a critical GFR threshold has not been investigated. Here, we use a survival random forest analysis for 15 years developed by Scheffner et al. where the graft survival was evaluated (these data are from an unpublished analysis which applied the same approach and cohort that was used in the study on patient graft survival from Scheffner et al^11^). Among one of the pivotal graft failure/success factors was first-year GFR.

Here, we model the patient graft survival probability as a sigmoidal function reading:

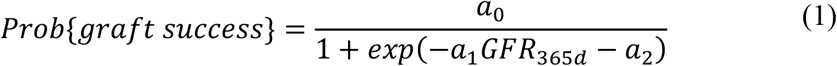

where *a*_*0*_ is the averaged maximum graft survival probability, and *α*_*1*_, and *α*_*2*_ arrange the shape of the function. The GFR values at the inflection point of the sigmoidal curve are considered as graft failure clinical thresholds for the different populations (*GFR*_*fail level*_).

Initially, we have calculated the survival probabilities of training cohort (N_1_) using a random forest survival model. Eq. 20 was fitted for all of the patients in the training group, regardless of rejection incidence. A safe range for failure critical threshold might be considered to be between 30 and 50 ml/min/1.73 m^2^ by comparing the likelihood of survival at 50% across all patient categories. All figures can be found in the Section 5 of SI (Figures S.6 – S.9).

### 2.3. Individualized prediction of critical GFR thresholds

An important task for our framework is the estimation of the individualized critical GFR threshold. The parameters of the Duffing oscillator (Eqs. 1-4) were estimated for all patients that belong to the group without rejection. As long as the conditions in Eqs. 5-8 were satisfied, the *a, b*, and *θ* parameters of the model were calculated by Eqs. 9-11. In turn, we drew the *λ* damping parameter from the empirical distribution found in Sec.3 SI.

In addition, from the prediction of GFR at 365 days and the use of the detailed mathematical analysis presented in section 2.3, the critical threshold can be estimated. Figure 1 contains the distribution of the personalized threshold *θ* in patients without rejection, one rejection, and multiple rejections for the training group. Statistical difference exists between the patients without rejection and patients with multiple rejections, and the patients with one and multiple rejections. The population with multiple rejections had smaller *θ* values in comparison with the other two patient groups. The average critical value for the three categories is 37±12 mL/min/1.73m^2^. In the following section, we will show the existence of such a critical GFR threshold with respect to graft survival.

**Figure 1:**
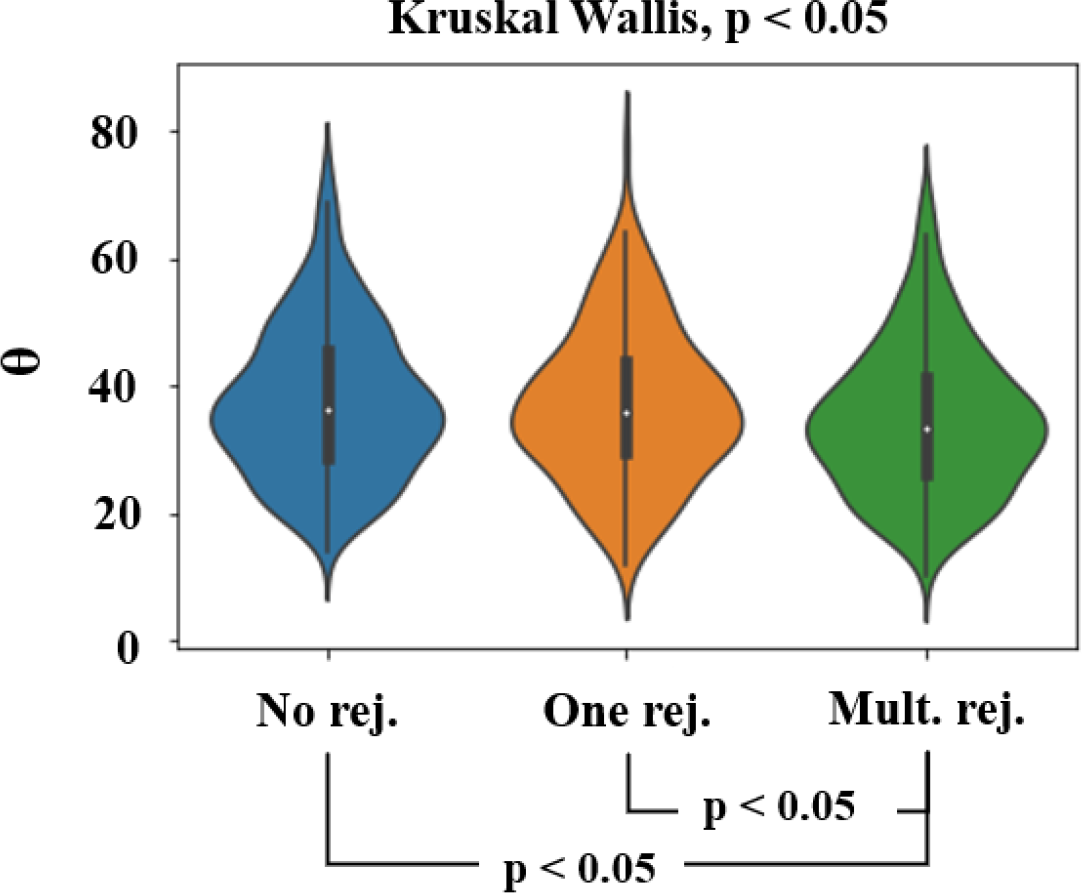
Distributions of critical GFR thresholds in patients with and without rejection in the training group.

### 2.4. Our algorithm satisfactorily predicts graft failure among all patient groups

Our regression analysis for predicting a patient’s GFR at 365 days postransplantation resulted in a prediction interval of ±18 units, across all three patient categories (Figures S2, S3, and S4 from supplementary material). The variation of GFR value between a minimum (*GFR*_min_) and maximum (*GFR*_max_) value led to the estimation of two more respective parameter sets of (*α, b, θ*) but the damping factor could remain uncertain. As stated above, the damping parameter λ was fitted from the non-rejection patient ensemble, including derivation of the corresponding probability distribution. Since for the parameter λ only the empirical distribution was known, we generated an ensemble of simulations for λ’s drawn from this distribution and a probability of graft failure was calculated (see Figure 2**Error! Reference source not found**.). This analysis was applied for the two populations (N_1_ and N_2_) predicting the graft risk for 365 days and 730 days post-transplantation.

**Figure 2:**
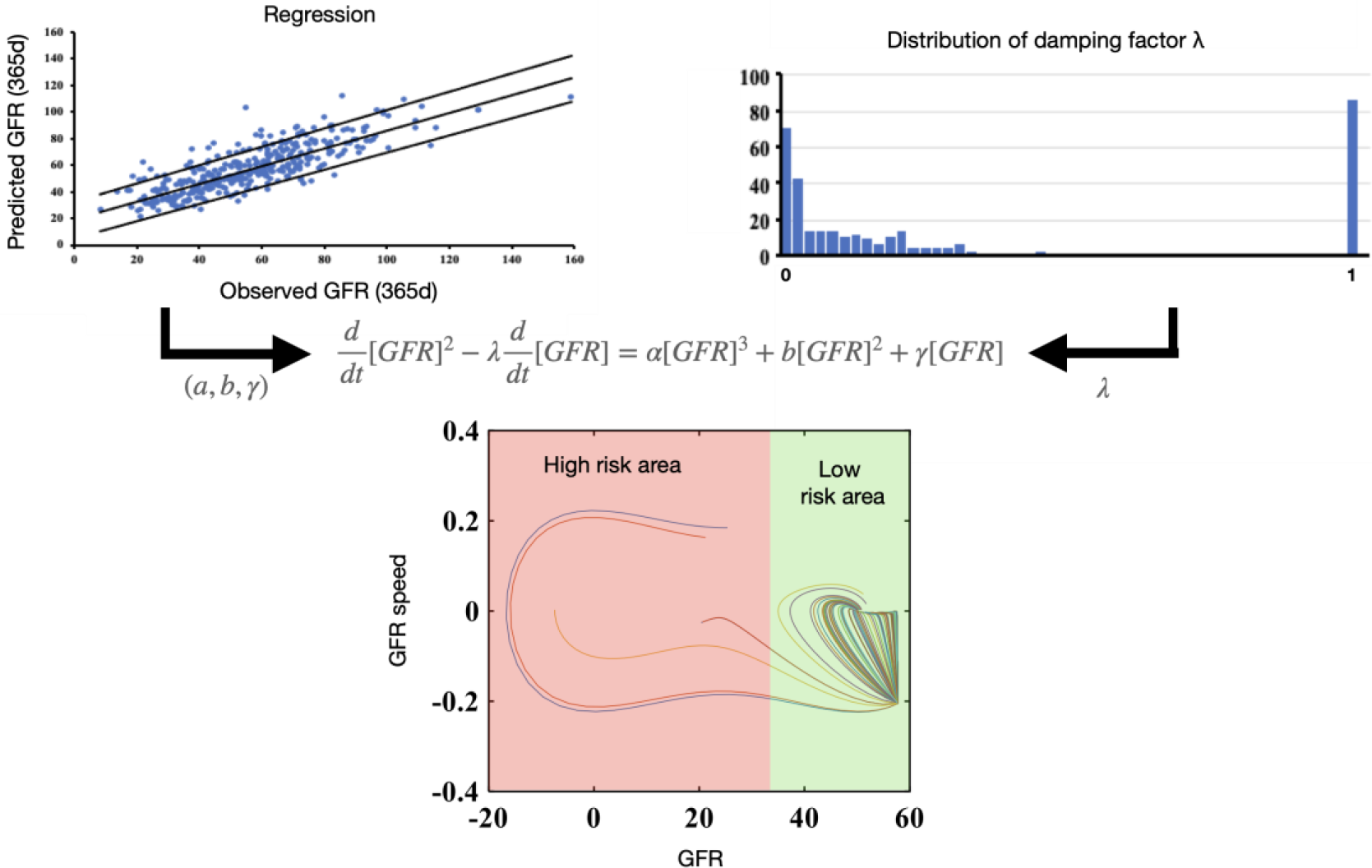
(A) The figure depicts the GFR dynamics in an example of a patient for a fixed (α, b, θ) parameter set, and (B) shows the 362 different values of λ drawn from the patient ensemble distribution. (C) The patient had a low probability of kidney failure, since most of the simulated trajectories landed above the critical threshold θ, implying a successful transplantation.

According to the steps discussed in section 2.4, we tested our risk assessment algorithm in the two groups N_1_ and N_2_. Starting from the training group N_1_ in Figure 3A and assuming a maximum predicted one-year GFR value, we obtained an over 80% area under the curve (AUC) across different GFR thresholds. To further validate the performance of our proposed methodology, the independent cohort group N_2_ was used, where all the computational procedures and the associated parameters remain the same. Figure 3A indicates that the N_2_ group has a similar AUC trend as the N_1_ group. Figure 3B shows the AUC in both groups. It is noticeable that their AUC values are all consistently above 0.8, reaching even values higher than 0.9, and the AUCs are close to each other between the two groups and across the categories of rejection/no rejection when the maximum threshold 50 mL/min/1.73 m^2^ was assumed.

**Figure 3:**
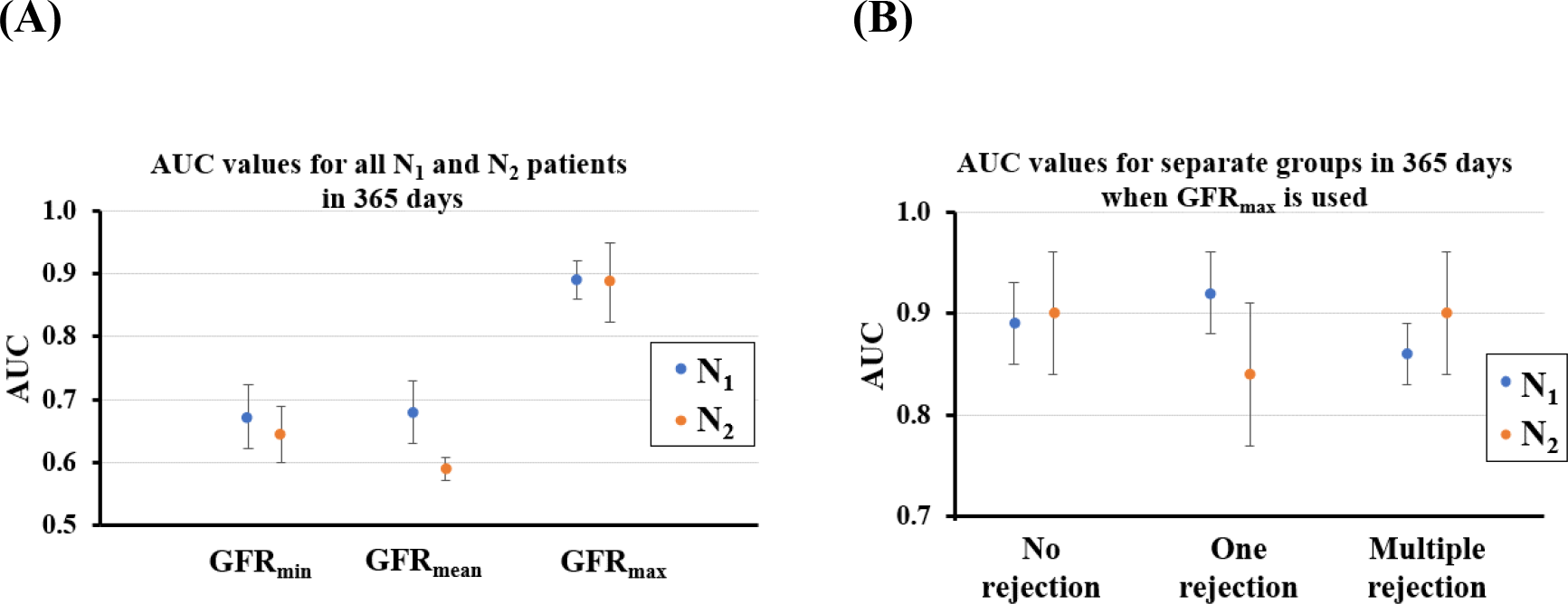
(A) Averaged AUC at 365 days with standard deviation in patients of both the training and validations groups (N_1_ and N_2_) over the different predicted GFR values in the critical thresholds ranging from 30-50 mL/min/1.73 m^2^. (B) averaged AUC with standard deviation of patient groups in N_1_ and N_2_ with and without rejection when critical thresholds ranged from 30-50 mL/min/1.73 m^2^.

Finally, by using the threshold concept, we examined if state-of-the-art classification methods such as XGBoost and Random Forest could outperform our algorithm. In particular, XGBoost was performing around 0.73 on average across patient groups and for different thresholds and Random Forests around 0.74 (for details see section 6 of SI). Therefore, the coupling of regression, mathematical modeling and uncertainty analysis had a better accuracy with respect to patient classification.

### 2.5. The graft risk prediction is robust in time

We attempted to explore the robustness of the prediction over time. A basic property of our mathematical model is that the functional state is a steady state, i.e. it is robust in time. This implies that the prediction of a patient classified as low risk using the GFR at 365 days should theoretically be consistent if applied to a later GFR measurement. We used the GFR values at 730 days and recalculated our predictions. Figure 4A depicts the AUC for different annual predicted GFR values at two years in both the training and validation groups. Figure 4B shows the AUC for both groups for patients with and without rejection, which is around 0.8. Indeed, the prediction performance was similarly good compared with the one-year prediction, suggesting that the prediction of graft risk failure is rather robust. Later time points of prediction were not tested as these may be affected by unforeseeable events, such as viral infections, that may deteriorate the graft function.

**Figure 4:**
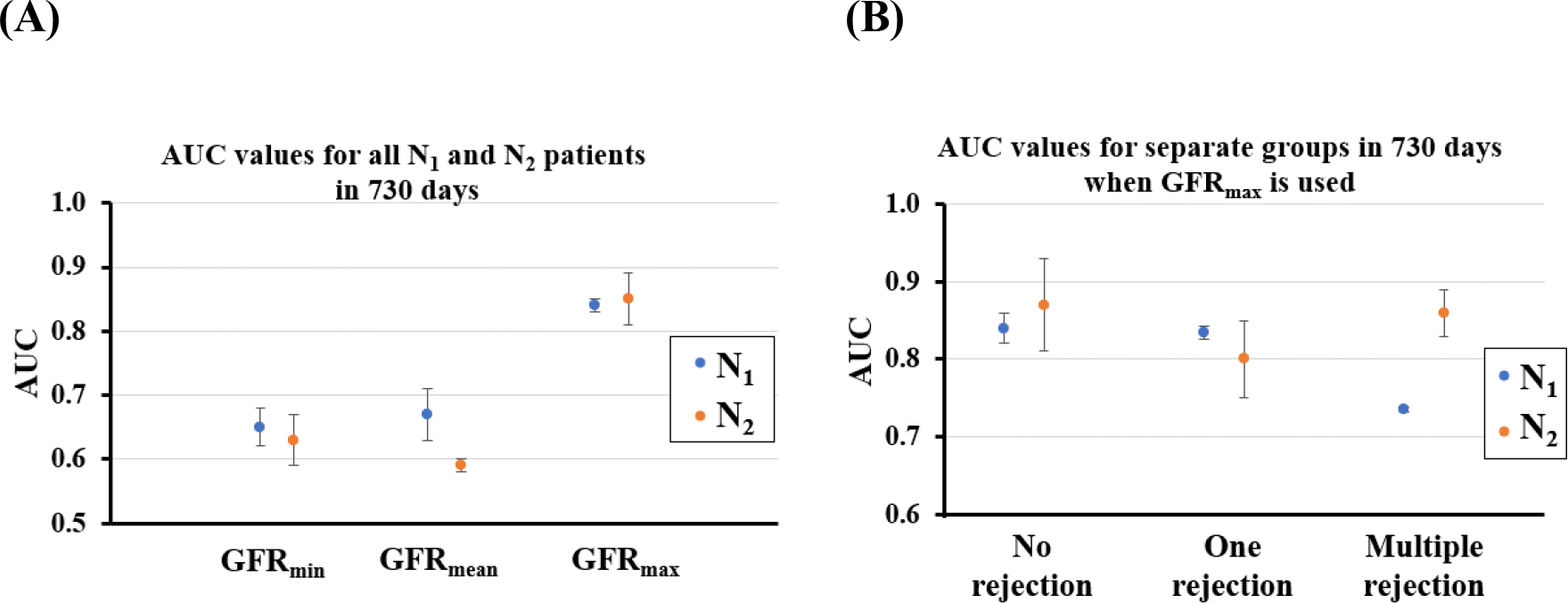
(A) Averaged AUC with standard deviation at 730 days post transplantation in patients of both the training and validation groups (N_1_ and N_2_) over the different predicted GFR values in the critical thresholds ranged from 30-50 mL/min/1.73 m^2^. (B) averaged AUC with standard deviation of the patient groups N_1_ and N_2_ with and without rejection when critical thresholds ranged from 30-50 mL/min/1.73 m^2^.

## 3. Discussion

This research introduces a framework for evaluating the graft function and risk of graft failure in kidney transplant patients using limited pre- and post-operative data. Utilizing state-of-the-art feature selection techniques, we have identified the key measurements that have the most significant impact on GFR after one year. The donor’s age, the highest GFR within 6 weeks after donation, and the percentage change in GFR at 100 days were found to be the most critical clinical parameters.

When we trained the regression model using data only from patients without rejection, we obtained reasonably predictive results for patients with and without rejection. However, we went further to enhance our model by developing a dynamic model capable of capturing GFR fluctuations and corresponding long-term GFR behavior. A critical assumption in our approach is the existence of a GFR threshold value that distinguishes successful grafts from failing ones. By utilizing survival functions from the N1 group, we were able to estimate this threshold.

It’s important to note that investigating the biological mechanisms underlying this threshold is beyond the scope of this paper. We used regression analysis to estimate model parameters, assuming it provides a robust estimate of individualized functional GFR (first-year value). Our proposed algorithm, trained on the N1 group without rejection, exhibited high AUC values for predicting graft risk at 365 and 730 days for all patients, both with and without rejection. This demonstrates the robustness of our method in distinguishing between low and high-risk patients regarding graft failure, even without providing exact GFR value predictions. Similar results were obtained when our algorithm was applied to the validation dataset (N2). Ultimately, we discovered that our method outperforms state-of-the-art classification methods, such as XGBoost and Random Forests, in predicting the fate of grafts.

While our current modeling approach describes individually expected GFR trajectories based on cases without rejection, our model can be expanded and refined to incorporate additional factors, such as different rejection phenotypes or other clinical complications mentioned earlier. This extension may help modulate the individual GFR trajectory. Additionally, variations in anti-rejection therapies and their responses can be incorporated into our proposed model. However, due to sample size limitations, we were unable to precisely determine the impact of different therapies. We are currently developing a stochastic model that incorporates time-varying factors to simulate and predict the temporal evolution of graft function more accurately. The introduction of stochasticity/noise will necessitate the use of Monte-Carlo simulations^12^ and Kramer’s theory to determine the graft’s fate. In addition, improved predictions could be made by applying regression analysis to real-time measurements more frequently and by updating the model parameters^13^.

Our approach has several limitations. The data used for model development and validation were collected from a single center, primarily serving patients of Caucasian origin. This homogeneity ensures consistency in treatment and patient responses but may reduce generalizability to ethnic groups and clinical practices. In addition, our modeling focused solely on rejection as the prime factor in graft injury and failure, omitting other significant factors like infections, drug toxicity, and relevant co-morbid conditions. Moreover, we did not further differentiate rejections in terms of severity, antibody- and T cell-mediated mechanisms, or responses to anti-rejection treatment. Finally, GFR formulae based on serum creatinine inherently possess errors and inaccuracies. Therefore, our approach may be more suitable for establishing an intra-individual relative range of variance in graft function rather than absolute GFR values.

Our current mathematical model describes the temporal GFR evolution in a somewhat phenomenological manner. The Duffing oscillator employs a third-order polynomial forcing term to represent the expected bistability between stable and failing grafts. A more biologically relevant model could enhance interpretability by explicitly modeling the involved pathophysiological processes, thus enabling the investigation of novel intervention strategies. As an example, graft injury of different causes may result in progressive organ fibrosis as suggested by our in vitro/in silico model (Setten at al^14^ which simulated the interplay of fibroblasts and macrophages under different oxygen stress and inflammatory conditions as function of a as a bistable state of injury and repair. This and the works of U. Alon in the topic^15,16^ could offer a solid starting point to develop biologically consistent alternative to the Duffing oscillator forcing term.

Our study serves as an example of combining dynamic modeling and machine learning to address a biomedical problem^17,18^. This is an emerging research field, with two distinct approaches: (i) data-driven derivation of dynamic models and (ii) data-assisted model predictions. In the former category are methods like SiNDy^19^ and Koopman operator learning approaches^20^. In the latter, the most prominent method is physics-informed neural networks^21^, which require knowledge of reliable dynamic models. In cases of model structural uncertainty, methods such as BAM^22^ are recommended. Finally, when dealing with noisy continuous measurements, data assimilation methods, with the Kalman filter^23^ being the most prominent, combine model predictions and statistical inference. Our approach relates to the specific problem of functional evolution of kidney grafts but should also be applicable to other settings of chronic disease with deteriotion of organ function.

## 4. Online Methods

### 4.1. Approach rationale

Our approach aims to calculate and estimate the individualized risk of graft failure for patients with a kidney graft. For this, we develop a mathematical model to predict the temporal GFR dynamics of a specific patient. The ingredients of the dynamic model are: (i) the oscillatory behavior of the GFR and (ii) the existence of two steady graft states. The former intends to model the fluctuations observed in GFR trajectories. The latter models the failed and the functional graft state, respectively. It is important to note that these states are assumed to be stable, which allows not only to make a temporal prediction but also to gain knowledge of the temporal persistence of the predicted state. This model involves a critical threshold GFR value (unstable) that separates the failed and functional graft states.

The challenge is the individualized parameter calibration of the model. In this regard, we employ the assumption that the first-year GFR value can be considered as a good proxy of the functional graft performance^3^, where the failed state can be assumed as null GFR. Using as a training set a cohort of patients, which represents the natural, undisturbed transplant course (patients without a rejection) as a training set, we employ a linear regression analysis to estimate the first-year GFR based on pre-operative patient data and two postoperative GFR measurements, namely the highest GFR measurement within six weeks after the transplantation and the value of GFR at one hundred days. Using these data, we estimate the individual patient’s model parameters (details in Materials & Methods).

The most important model parameter is the GFR threshold value. GFR values above it imply a successful transplantation with a stable functioning graft, whereas values below indicate a high risk of graft failure. Using uncertainty analysis techniques, we calculate a probability for the risk of graft failure. Finally, we validate our predictions in two independent data sets, consisting of patients with no, one, or multiple rejections by using a small number of inputs. The model was calibrated only for patients without rejection in order to avoid any data leakage. Our strategy is succinctly described in Figure 5.

**Figure 5:**
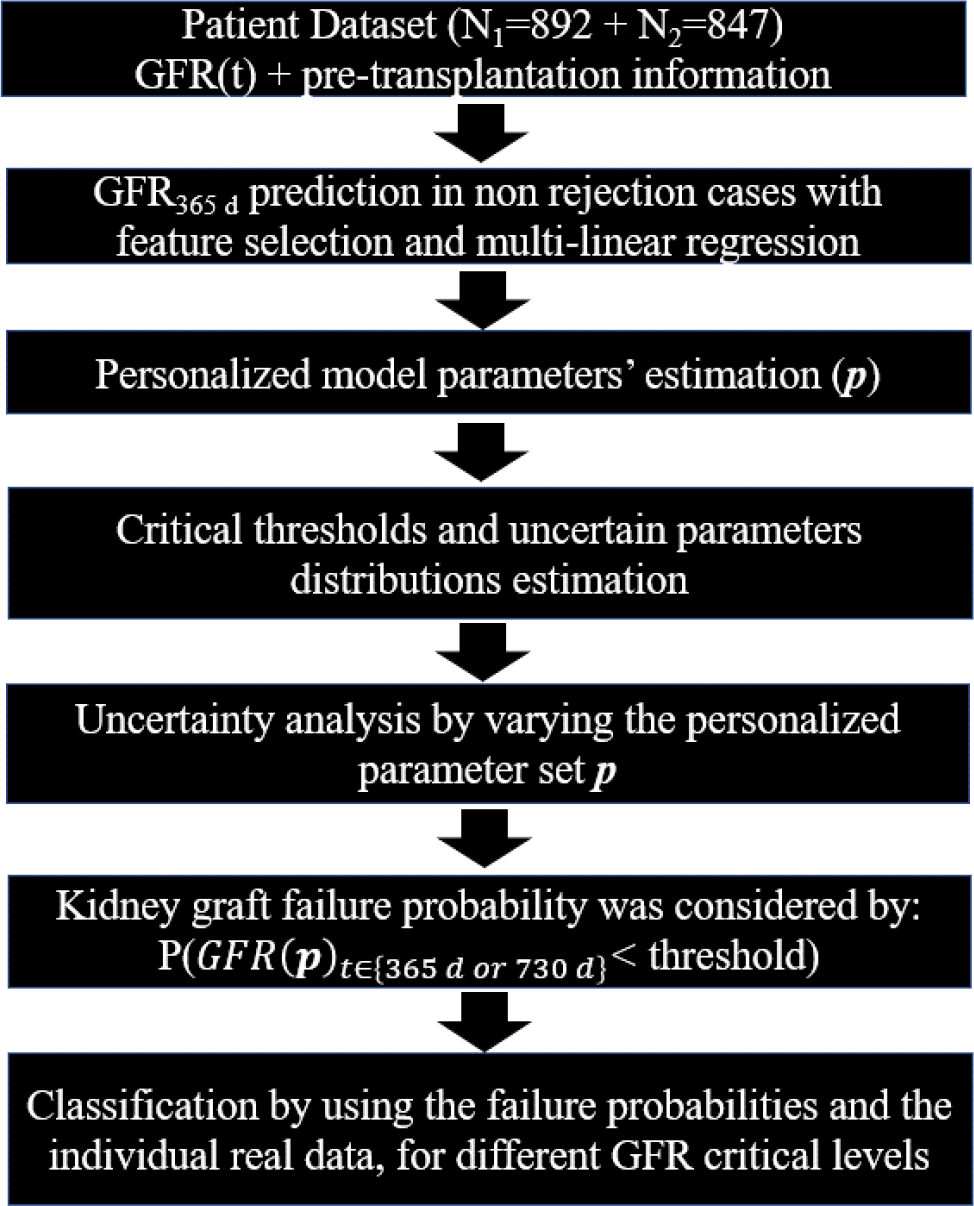
Steps followed from experimental measurements to the classification of the patients.

### 4.2. Clinical data

Data was taken from adult patients who received a kidney transplant alone or combined with another solid organ between 2000 and 2015 at Hannover Medical School and participated in our protocol biopsy program. Data collection and analysis were performed with the informed consent of the patients and with the approval of the institutional review board (no 2765). Two different patient sets were used in this study. The training group N_1_ and the validation cohort N_2_ consisted of 892 and 847 patients, respectively. In the training group, 362 patients had no rejection, 187 patients had one rejection, and 343 patients had multiple rejections. In the validation cohort, 426 patients had no rejection, 159 patients had one rejection, and 262 patients had multiple rejections. Protocol biopsies were collected six weeks, 3, and 6 months after transplantation. Also, data collection was performed before and at transplantation. Data of additional biopsies for cause, performed upon unexplained graft deterioration (at any time), were also recorded. Biopsies were evaluated according to the Banff classification valid at the time of biopsy^24^.

Anti-thymocyte globulin was used in sensitized patients and for combined kidney/ pancreas transplantations. The standard maintenance therapy consisted of a calcineurin inhibitor, mycophenolate mofetil, and prednisolone. Until the end of 2004, patients with low immunological risk (i.e., first kidney transplant from deceased donors in patients without panel-reactive antibodies) received a dual maintenance therapy with a calcineurin inhibitor and prednisolone. Tacrolimus gradually replaced cyclosporine A over the years.

Acute T cell-mediated rejections (TCMR), including borderline cases were treated with steroid boli, with the exception of subclinical borderline cases in protocol biopsies, defined by an increase in serum creatinine over baseline by <20% at biopsy. In addition, mycophenolate mofetil was added in patients on a dual immunosuppressive maintenance therapy. Patients with TCMR occurring at 6 months or later or with vascular TCMR at any time point were switched from cyclosporine A to tacrolimus. Acute antibody-mediated rejection (ABMR) was treated with steroid boli, plasma exchange, rituximab, immune globulins, and a switch to or increase in tacrolimus when full histopathomorphological criteria of the rejection were present (with or without donor-specific antibodies), interstitial fibrosis and tubular atrophy was <25%, and baseline estimated glomerular filtration rate (eGFR) before the rejection was >25 ml/min. In all other cases, individual treatment decisions were made, mostly consisting of steroid boli, immune globulins, a switch to or increase in tacrolimus, or no treatment in cases with minor findings of glomerulitis and peritubular capillaritis. No standardized treatment was defined for BK polyomavirus nephropathy or other findings (e.g., acute tubular injury, tubulointerstitial fibrosis/tubular atrophy, glomerulonephritis).

The glomerular filtration rate was calculated with the Cockcroft and Gault formula, and then GFR was estimated in units mL/min/1.73 m^2^ body surface. The criteria for delayed graft function were <500 mL urine within the first 24 hours post-transplantation and/or dialysis treatment necessary within the first week

### 4.3 Prediction of first-year graft function

Initially, only patients without rejection from the training group were used for feature selection and multilinear regression to predict the GFR at 365 d. The multilinear regression model included pre-transplantation data and modellable parameters, including 1) the age of the patient, 2) the sex of the patient, 3) the donor’s age, 4) the donor’s sex, 5) deceased donor/living donor graft, 6) cold ischemia time (hours), 7) presence or absence of delayed graft function, 8) HLA-mismatch at locus A, 9) HLA-mismatch at locus B, 10) HLA-mismatch at locus DR, 11) level of pre-formed antibodies at transplantation, 12) blood transfusions before transplantation, 13) the number of pregnancies before transplantation, 14) the number of previous transplantations, 15) the highest GFR achieved within six weeks after transplantation, and 16) the percentage of change between the highest GFR within six weeks and the GFR at 100 d. Since GFR measurement times were non-uniform i.e., GFR was sampled at various post-operative times, we have interpolated the GFR values for every 10 days for the first 100 days.

A sequential forward selection was performed using the Akaike criterion (AIC), the Bayesian information criterion (BIC), and the adjusted R squared for the best model selection^25^. The regression coefficients represented the importance of the underlying features. The derived multilinear model was assumed to be applicable to all other patient groups with one rejection and multiple rejections.

### 4.4. Mathematical modeling of GFR evolution

GFR dynamics of patients are typically characterized by a growth phase in a short period after transplantation^3^. In individual patients, the dynamics in GFR evolution may show weak and damped oscillations toward a specific GFR value or a decreasing trend that may result in graft failure (see Figure 6).

**Figure 6:**
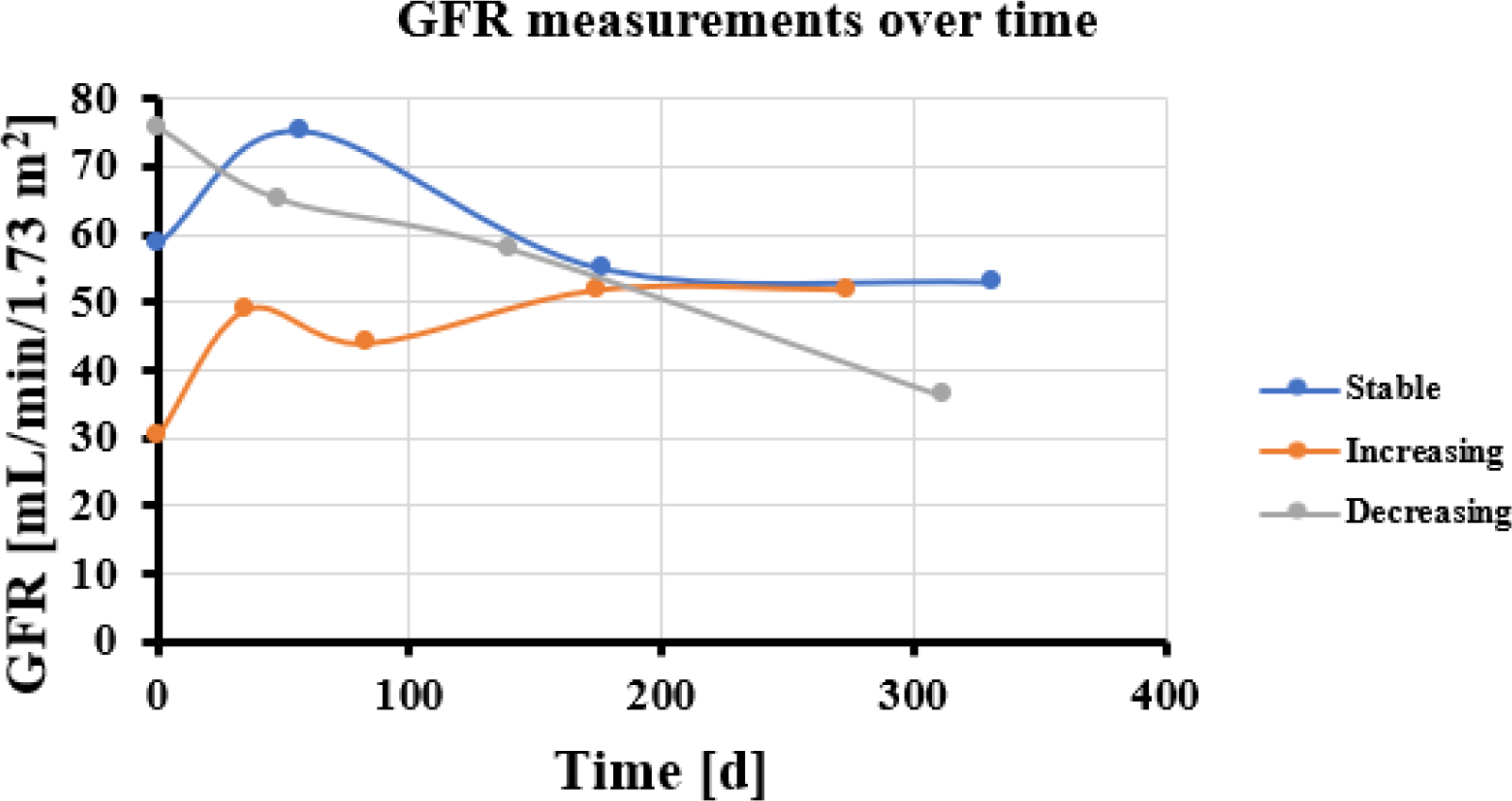
Examples of different graft function time courses of individual patients.

We modeled this GFR oscillatory behavior with a phenomenological Duffing oscillator (Figure 7A). Our selection is justified by the existence of two equilibria, i.e., the failed and the functional kidney state (Figure 7A). These equilibria are typically separated by a GFR threshold *θ* which is mathematically characterized asa saddle-node (unstable). Finally, the functional steady-state is estimated as the predicted first-year GFR for each patient using a regression approach. The failed graft steady-state is assumed as GFR=0 mL/min/1.73 m^2^. The Duffing oscillator model is described by the following ordinary differential equations (ODEs):

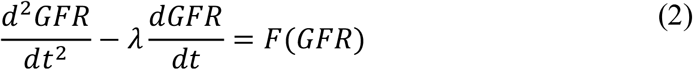

Where specifically we have:

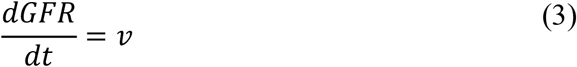

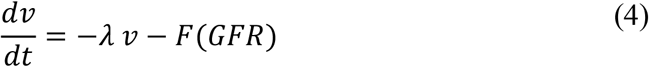

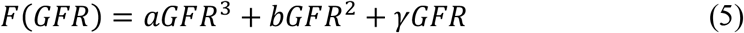

The variable *v(t)* is the GFR speed (rate of change), *GFR*(t) denotes the temporal evolution of GFR (ml/min/1.73 m^2^ body surface) and *dGFR*/*dt* denotes the time-derivative (speed) of the GFR. The parameter *λ* controls the amount of damping. The right hand side is a “forcing” term, which phenomenologically lumps the underlying biological processes responsible for GFR production. The simplest choice of function *F* is assumed to be given by a third-order polynomial. *α, b*, and *γ* are model parameters which are estimated individually.

**Figure 7:**
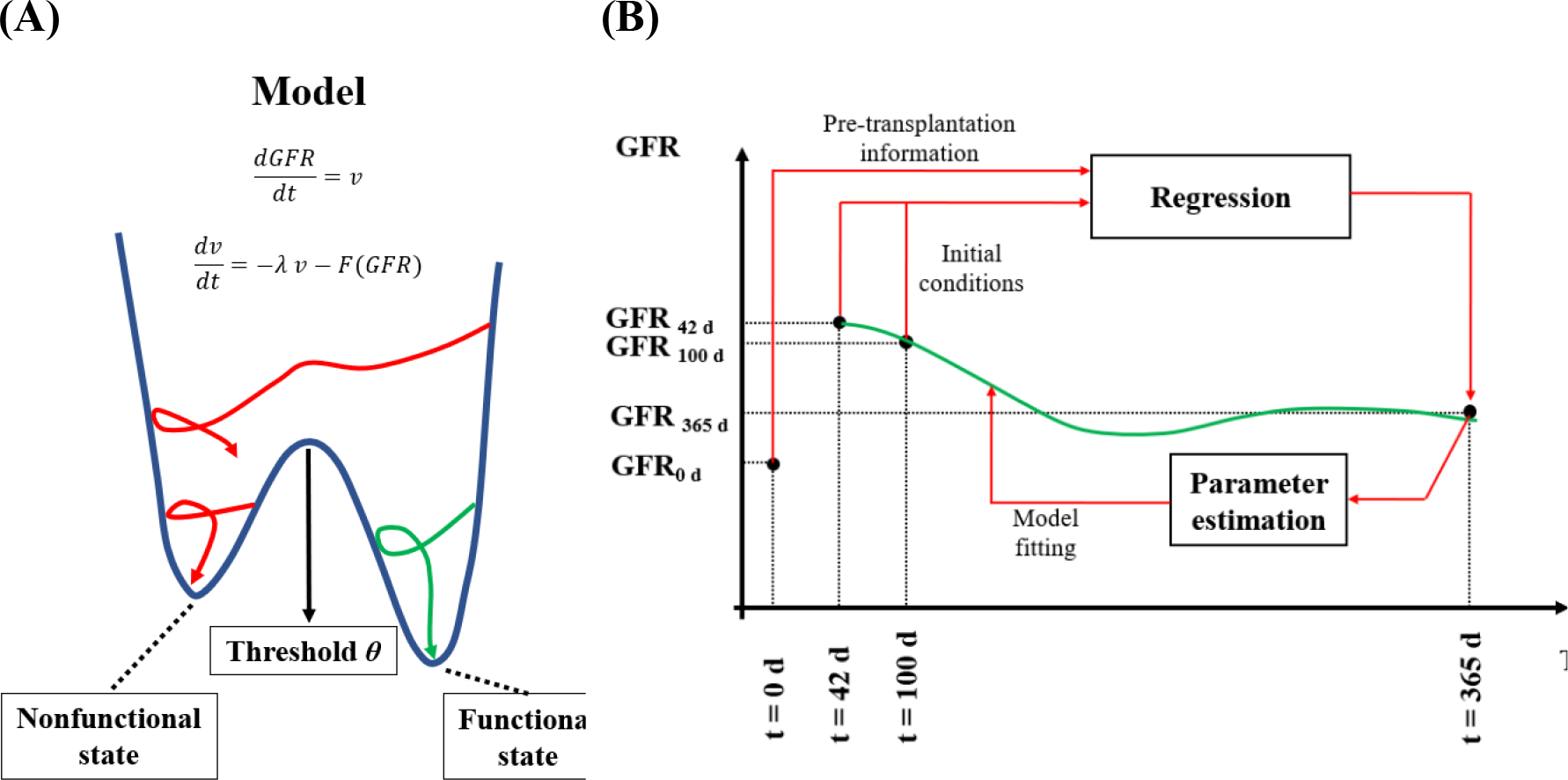
(A) Patient trajectories illustrated on the Duffing oscillator potential. (B) Model calibration approach for our GFR dynamic model combining the contribution of 6 weeks-best GFR (42 days) and 100 days GFR by regression analysis.

The steady states of the Duffing oscillator are related to GFR values of the failed, functional and the threshold (saddle-node) state. To explicitly calculate the afore-mentioned steady states, regarding the threshold *θ*, we set:

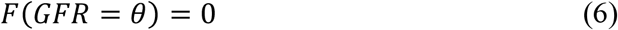

Which leads to:

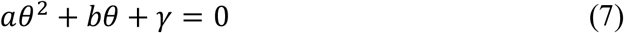

And we also set at 365 days

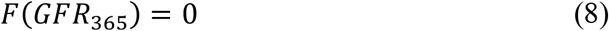

Where *GFR*_365_ is the GFR at 365 d. Eq. 7 leads to:

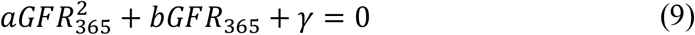

Combining Eqs. 6 and 8, with *γ* estimated parameter values, *a, b*, and *θ* parameters are approximated to:

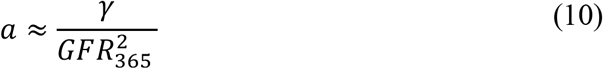

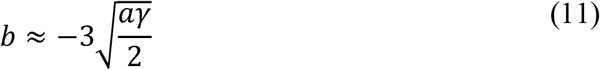

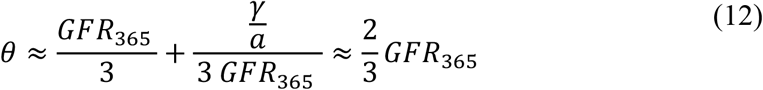

Section 1 of the Supplementary Material includes details for all the steps needed for the parameter approximations.

The GFR value at 365 days (*GFR*_365_) was predicted for all patients in the groups with and without rejection by the regression model trained by the patient cohort without any rejection episode (Section 2 of the SI including Figures S.1.-S.4. and Tables S.1. and S.2.). When patients with relevant complications which are known to affect the graft (rejection, BK virus nephropathy, glomerulonephritis, urinary tract infections, other severe infections and other severe extra-renal diseases) were excluded the annual rate of change was 1.58 ml/min/1.73 m^2^ (Table S.3 in the Section 3 of SI). The whole cohort of patients had an average annual GFR loss of 2.07 ml/min/1.73 m^2^ (Table S.4 in the Section 3 of SI). Thus, *γ* could be estimated equal to 10^-4^ d^-2^ (Section 3 in the SI).

With the individualized parameters calibrated as described above and starting with the GFR at six weeks, the initial condition of the model (1) can be set for each individual patient. Then, using the patient cohort without rejection as a training set again, the *λ* parameter was estimated individually from the clinical measurements with the particle swarm optimizer algorithm (PSO)^26^. PSO belongs to stochastic, population-based computer algorithms. The main concept of this method is that each parameter which is considered as a particle, is a design point and is available to move in multi-dimensional space for finding the best solution ^27^. Thus, the time-evolution of the GFR can be simulated for each patient, when a *λ* is selected from the obtained distribution (details in Section 4 of SI). This procedure of parameter calibration is shown in Figure 6B for a hypothetical patient. Finally, having all model parameters estimated, a personalized GFR threshold can be approximated from Eq. 11.

### 4.5. Individualized graft failure risk assessment

Our goal is to use the calibrated model to calculate the individual graft failure risk. Initially, a range for the predicted GFR value at 365 d was defined, taking into account the regression mean square error for all different patient groups with and without rejection. According to the individualized minimum, mean, and maximum GFR predictions from the regression model, the parameters *α, b*, and *θ* were calibrated. The parameter *λ* was drawn from an empirical distribution calculated from the patient cohort without rejection, as this population’s GFR values were less affected by anti-rejection treatments.

Each *i*-th patient GFR was simulated n times for the 3 different 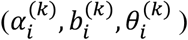 parameter sets for *t*_*1*_ = 365 d, or *t*_*2*_ = 730 d. An ensemble of *n* simulations was defined, where for each *j*-th simulation a *λ*_*j*_ value was drawn from the corresponding distribution. A simulated graft was considered failed, when the GFR at the end of the simulation was less than the threshold *θ*_*i*_, i.e., *GFR*_*i*_(t)<*θ*_*i*_ (Figure 7). Finally, a failure probability was calculated as the fraction of “failed” simulated grafts. More precisely, the steps are given in the box below.

- A parameter set 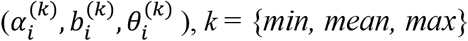is estimated for each *i*-th patient.
- For every *t* ∈{*t*_1_ = 365 d, *t*_2_ = 730 d}, and *λ*_*j*_ ∈{ *λ*_1_, …, *λ*_n_}, where n number of samples drawn, the 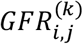 (*t*) is estimated
- Assumed indicator function *I*, where:

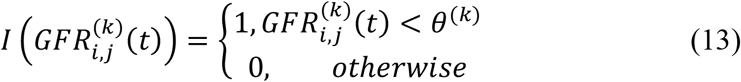
- The probability of failure is calculated as:

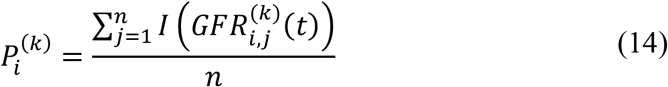

### 4.6. Validation process

In turn, the receiver operating characteristic curve (ROC) was created by setting a threshold probability (*p*_thr_) range from 0 to 1, and comparing the actual values with the real kidney transplantation outcomes. More precisely, regarding the low and high risk for each patient at 365 d and at 730 d, the true success, the false failure, the false success, and the true failure were estimated as follows:

- True success (right low risk patient prediction):

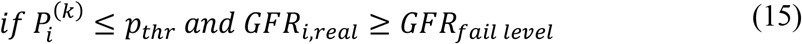
- The false failure (false high risk patient prediction):

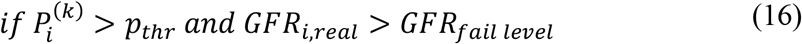
- The false success (false low risk patient prediction):

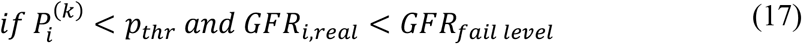
- The true failure (true high risk patient prediction):

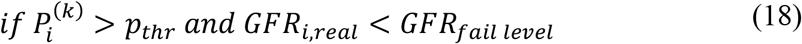

The ROC was ploted bases on true success and false success rates, which are equal:

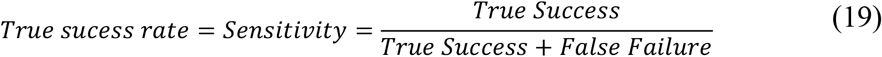

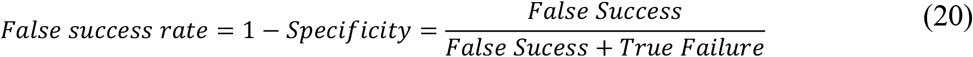

The corresponding area under the curve (AUC) was calculated for each ROC in order to select the best parameter set that represents the individualized risk.

## Data Availability

All data produced in the present study are available upon reasonable request to the authors

## Acknowledgments

S. Kadah and N.S. Heinz from the Hannover Medical School (MHH) assisted with data management. H.H. have received funding from the Volkswagenstiftung and the “Life?” program (96732). H.H. has received funding from the Bundes Ministerium für Bildung und Forschung under the grant agreement No. 031L0237C (MiEDGE project/ERACOSYSMED). Finally, H.H. acknowledges the funding of the FSU grant 2021-2023 grant from Khalifa University. WG has received funding under the frame of ERACoSysMed-2, the ERA-Net for Systems Medicine in clinical research and medical practice (project ROCKET, JTC2_29).

## Data and code availability

The code with the data can be made available upon reasonable requesst.

## Ethics approval and consent to participate

Data collection and analysis were performed with prior informed consent of the patients and with the approval of the Hannover Medical School Ethics review board (no 2765) that decides on all patient studies, according to national laws and based on the Declaration of Helsinki and regarding transplant patients, on the Declaration of Istanbul.

## Author Contributions

Conceptualization, G.W. and H.H..; methodology, S.S., A.R. and H.H.; software, S.S. I.S. A.R. and H.H.; validation, S.S., H.H and G.W.; formal analysis, S.S., I. S., A.R., and H.H.; investigation, S. S., I.S., A.R., H.H., and G.W.; resources, I.S. and G.W.; data curation, S.S., A.R., I.S. and G.W.; writing—original draft preparation, S.S. I.S., A.R., H.H. and G.W.; writing—review and editing, S.S., I.H., A.R. H.H. and G.W.; visualization, S.S., I.H., A.R. H.H. and G.W.; supervision, H.H. and G.W.; project administration, H.H., and G.W.;

## References

1. Ng, K., Kartoun, U., Stavropoulos, H., Zambrano, J. A. & Tang, P. C. Personalized treatment options for chronic diseases using precision cohort analytics. Sci Rep 11, (2021).

2. Bakris, G. L. et al. Preserving renal function in adults with hypertension and diabetes: a consensus approach. National Kidney Foundation Hypertension and Diabetes Executive Committees Working Group. Am J Kidney Dis 36, 646–61 (2000).

3. Kettler, B. et al. Reply to Sabah et al. Transplant International 32, 1341–1342 (2019).

4. National Institute of Diabetes and Digestive and Kidney Diseases (U.S.). Division of Kidney Urologic and Hematologic Diseases., Urban Institute. Renal Research Program., University of Michigan. & USRDS Coordinating Center. United States Renal Data System. Annual Data Report. v. (2008).

5. Humar, A. & Matas, A. J. Surgical complications after kidney transplantation. Semin Dial 18, 505–510 (2005).

6. Matz, M. et al. Identification of T cell-mediated vascular rejection after kidney transplantation by the combined measurement of 5 specific MicroRNAs in blood. Transplantation 100, 898–907 (2016).

7. Luque, S. et al. Value of monitoring circulating donor-reactive memory B cells to characterize antibody-mediated rejection after kidney transplantation. American Journal of Transplantation 19, 368–380 (2019).

8. Van Den Brand, J. A. J. G. et al. Predicting kidney failure from longitudinal kidney function trajectory: A comparison of models. PLoS One 14, (2019).

9. Armero, C., Forte, A., Perpiñán, H., Sanahuja, M. J. & Agustí, S. Bayesian joint modeling for assessing the progression of chronic kidney disease in children. Stat Methods Med Res 27, 298–311 (2018).

10. Van Loon, E. et al. Forecasting of Patient-Specific Kidney Transplant Function with a Sequence-to-Sequence Deep Learning Model. JAMA Netw Open 4, (2021).

11. Scheffner, I., Gietzelt, M., Abeling, T., Marschollek, M. & Gwinner, W. Patient Survival after Kidney Transplantation: Important Role of Graft-sustaining Factors as Determined by Predictive Modeling Using Random Survival Forest Analysis. Transplantation 1095–1107 (2020) doi:10.1097/TP.0000000000002922.

12. Cantisán, J., Seoane, J. M. & Sanjuán, M. A. F. Stochastic resetting in the Kramers problem: A Monte Carlo approach. Chaos Solitons Fractals 152, 111342 (2021).

13. Solomon, R. & Goldstein, S. Real-Time measurement of glomerular filtration rate. Curr Opin Crit Care 23, 470–474 (2017).

14. Setten, E. et al. Understanding fibrosis pathogenesis via modeling macrophage-fibroblast interplay in immune-metabolic context. Nature Communications 2022 13:1 13, 1–22 (2022).

15. Adler, M. et al. Principles of Cell Circuits for Tissue Repair and Fibrosis. iScience 23, (2020).

16. Zhou, X. et al. Circuit Design Features of a Stable Two-Cell System. Cell 172, 744–757.e17 (2018).

17. Hatzikirou, H. Combining dynamic modeling with machine learning can be the key for the integration of mathematical and clinical oncology: Comment on ‘Improving cancer treatments via dynamical biophysical models’ by M. Kuznetsov, J. Clairambault, V. Volpert. Phys Life Rev 40, 1–2 (2022).

18. Karniadakis, G. E. et al. Physics-informed machine learning. Nature Reviews Physics 2021 3:6 3, 422–440 (2021).

19. Rudy, S. H., Brunton, S. L., Proctor, J. L. & Kutz, J. N. Data-driven discovery of partial differential equations. Sci Adv 3, (2017).

20. Williams, M. O., Kevrekidis, I. G. & Rowley, C. W. A Data–Driven Approximation of the Koopman Operator: Extending Dynamic Mode Decomposition. J Nonlinear Sci 25, 1307–1346 (2015).

21. Raissi, M., Perdikaris, P. & Karniadakis, G. E. Physics-informed neural networks: A deep learning framework for solving forward and inverse problems involving nonlinear partial differential equations. J Comput Phys 378, 686–707 (2019).

22. Mascheroni, P., Savvopoulos, S., Alfonso, J. C. L., Meyer-Hermann, M. & Hatzikirou, H. Improving personalized tumor growth predictions using a Bayesian combination of mechanistic modeling and machine learning. Communications Medicine 2021 1:1 1, 1–14 (2021).

23. Oikonomou, V. P., Tzallas, A. T., Konitsiotis, S., Tsalikakis, D. G. & Fotiadis, D. I. The Use of Kalman Filter in Biomedical Signal Processing. in Kalman Filter (eds. Moreno, V. M. & Pigazo, A.) (IntechOpen, 2009). doi:10.5772/6805.

24. Loupy, A. et al. The Banff 2019 Kidney Meeting Report (I): Updates on and clarification of criteria for T cell– and antibody-mediated rejection. American Journal of Transplantation 20, 2318–2331 (2020).

25. Marcano-Cedeño, A., Quintanilla-Domínguez, J., Cortina-Januchs, M. G. & Andina, D. Feature selection using Sequential Forward Selection and classification applying Artificial Metaplasticity Neural Network. IECON Proceedings (Industrial Electronics Conference) 2845–2850 (2010) doi:10.1109/IECON.2010.5675075.

26. Bonyadi, M. R. & Michalewicz, Z. Particle swarm optimization for single objective continuous space problems: A review. Evol Comput 25, 1–54 (2017).

27. Jansen, P. W. & Perez, R. E. Constrained structural design optimization via a parallel augmented Lagrangian particle swarm optimization approach. Comput Struct 89, 1352–1366 (2011).

