## Supplementary Material for "A model-driven machine learning approach for personalized kidney graft risk prediction"

a. Department of Mathematics, Khalifa University, Abu Dhabi, United Arab Emirates.
b. Department of Nephrology and Hypertension, Hannover Medical School, Hannover, Germany.
c. Charité-Universitätsmedizin Berlin, corporate member of Freie Universität Berlin, Humboldt-Universität, Berlin, Germany

*These authors share the first authorship,
+ These authors share the last authorship

**Section 1: Parameters of the model.**

The duffing oscillator model is given by:

$$\frac{d^2GFR}{dt^2} - \lambda \frac{dGFR}{dt} = f(GFR) \qquad \text{(S. 1)}$$

Where:

$$f(GFR) = a\, GFR^3 + b\, GFT^2 + \gamma\, GFR + \delta \qquad \text{(S. 2)}$$

Which derives:

$$\frac{dGFR}{dt} = u \qquad \text{(S. 3)}$$

$$\frac{du}{dt} = -\lambda\, u - f(GFR) \qquad \text{(S. 4)}$$

If $c$=$GFR$ at 365 d, and $\theta$ is the threshold, then at steady-state, the model should satisfy equations S. 5, S. 6, and S. 7.

$$f(0) = 0 \;\; => \;\; \delta = 0 \qquad \text{(S. 5)}$$

$$\begin{gathered} f(c)=0 => ac^3 + bc^2 + \gamma c = 0 \Leftrightarrow \\ \Leftrightarrow ac^2 + bc + \gamma = 0 \Leftrightarrow \\ \Leftrightarrow a = -\frac{bc + \gamma}{c^2} \end{gathered} \qquad \text{(S. 6)}$$

$$f(\theta) = 0 \;\Leftrightarrow\; a\theta^2 + b\theta + \gamma = 0 \qquad \text{(S. 7)}$$

At GFR at 365 days:

$$\frac{df}{dx} < 0 \Leftrightarrow 3\alpha c^2 + 2bc + \gamma < 0 \qquad \text{(S. 8)}$$

At the threshold $\theta$:

$$\frac{df}{dx} > 0 \Leftrightarrow 3\alpha\theta^2 + 2b\theta + \gamma > 0 \qquad \text{(S. 9)}$$

Also, the curvature changes at the threshold

$$\begin{gathered} \frac{d^2f}{dx^2} = 0 \Leftrightarrow 6\alpha\theta + 2b = 0 \Leftrightarrow \\ \Leftrightarrow \theta = -\frac{b}{3\alpha} \end{gathered} \qquad \text{(S. 10)}$$

From equations S. 7 and S. 10:

$$\begin{gathered} \frac{ab^2}{9\alpha^2} - \frac{b^2}{3\alpha} + \gamma = 0 \Leftrightarrow \\ \Leftrightarrow b^2 - 3b^2 + 9\alpha\gamma = 0 \Leftrightarrow \\ \Leftrightarrow b = \pm 3\sqrt{\frac{\alpha\gamma}{2}} \end{gathered} \qquad \text{(S. 11)}$$

In equations S. 2 and S. 5, $\gamma$ should be positive as it represents the decay of *GFR* in the patients. Thus, from equation S. 11, $\alpha$ is positive. Based on equations S. 10 and S. 11, $b = -3\sqrt{\frac{\alpha\gamma}{2}}$ because $\theta>0$.

$\theta$ then equals:

$$\begin{gathered} \theta = -\frac{b}{3\alpha} = -\frac{-3\sqrt{\frac{\alpha\gamma}{2}}}{3a} \Leftrightarrow \\ \Leftrightarrow \theta = \sqrt{\frac{\gamma}{2\alpha}} \end{gathered} \qquad \text{(S. 12)}$$

From equation S. 6, by replacing *b*, $\gamma$ as a function of *c* and *a* can be approximated by:

$$\begin{gathered} ac^2 - 3\sqrt{\frac{\alpha\gamma}{2}}c + \gamma = 0 \Leftrightarrow \\ \Leftrightarrow ac^2 - \frac{3}{\sqrt{2}}c\sqrt{a}\sqrt{\gamma} + \gamma = 0 \Leftrightarrow \\ \Leftrightarrow ac^2 - 2c\sqrt{a}\sqrt{\gamma} + \gamma + 2c\sqrt{a}\sqrt{\gamma} - \frac{3}{\sqrt{2}}c\sqrt{a}\sqrt{\gamma} = 0 \Leftrightarrow \\ \Leftrightarrow \left(c\sqrt{a} - \sqrt{\gamma}\right)^2 + \frac{2\sqrt{2} - 3}{\sqrt{2}}c\sqrt{a}\sqrt{\gamma} = 0 \end{gathered} \qquad \text{(S. 13)}$$

The second term of equation S. 13 can be deleted as $\alpha$ and $\gamma$ are quite small. Therefore, $\gamma$ can be approximated by:

$$\gamma \approx \alpha c^2 \tag{S. 14}$$

From equations S. 6 and S. 10:

$$c^2 - 3\theta c + \frac{\gamma}{\alpha} = 0 \Leftrightarrow$$
$$\Leftrightarrow \theta = \frac{c^2 + \gamma/\alpha}{3c} \tag{S. 15}$$

Using equation S. 14 to S. 13, $\theta$ can also be approximated from measurable $c$ by:

$$\theta \approx \frac{2c}{3} \tag{S. 16}$$

**Section 2: Feature selection and multi-linear regression.**

Patients without rejection from the training set were used for feature selection and prediction of GFR after 365 days postransplantation. The inputs were 1) patient age, 2) patient sex, 3) donor's age, 4) donor's sex, 5) deceased donor vs. living donor 6) cold ischemia time (hours), 7) initial function vs. delayed function of the graft, 8) HLA-mismatch at locus A, 9) HLA-mismatch at locus B, 10) HLA-mismatch at locus DR, 11) panel reactive antibodies (%), 12) blood transfusions before transplantation, 13) number of pregnancies, 14) number of previous transplantations, 15) highest GFR within six weeks after transplantation (ml/min/1.73 $m^2$), and 16) percentage change between the highest GFR within six weeks and the GFR at 100 d. All inputs were used in the AIC, BIC and adjusted $R^2$ for feature selection in Python. The results of the feature selection are shown in Figure S. 1.

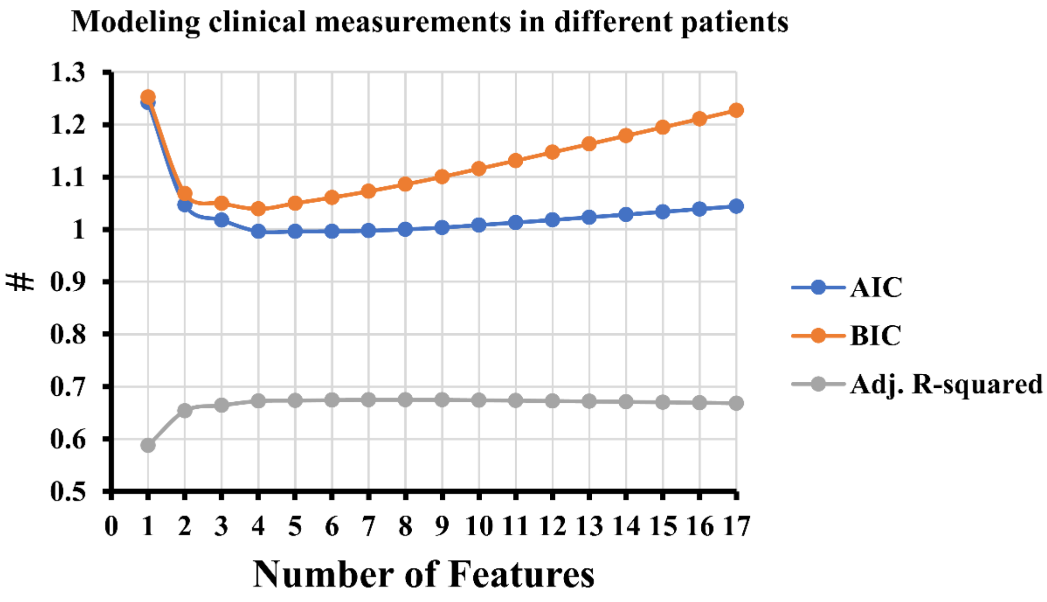


**Figure S. 1:** AIC, BIC and adjusted $R^2$ values over the number of features.

Table S. 1 contains the number of the features and the index of the features for the first five numbers.

**Table S. 1:** Important features in the first five feature steps

| Number of features | Index of the features |
|---|---|
| 1 | $GFR_{in}$[1] |
| 2 | $GFR_{in}$, % of GFR change |
| 3 | $GFR_{in}$, % of GFR change, donor's age |
| 4 | $GFR_{in}$, % of GFR change, donor's age, blood transfusions before transplantation |
| 5 | $GFR_{in}$, % of GFR change, donor's age, blood transfusions before transplantation, HLA-mismatch at locus B |

[1] $GFR_{in}$: Highest GFR within the first 6 weeks posttransplantation

According to Figure S. 1 and Table S. 1, the selected number of features was three as it showed a good performance, and each criterion value varied insignificantly when the number of features increased.  Thus, the most important features were donor's age ($x_1$), the highest GFR within six weeks after transplantation ($x_2$), and the percentage change between the highest GFR within six weeks and the GFR at 100 d ($x_3$).

The corresponding coefficients of the features with reference to multi-linear regression, and the intercept for predicting the GFR 365 days posttransplantation, are shown in Table S. 2.

**Table S. 2:** Ordinary least squares regression results

| **Feature** | **Value** | **Std err** | **t** | **P>\|t\|** | **95% CI** |
|---|---|---|---|---|---|
| Intercept | 19.5974 | 3.608 | 5.431 | 0.000 | [12.501 26.694] |
| $x_1$ | -0.1647 | 0.047 | -3.513 | 0.001 | [-0.257 -0.072] |
| $x_2$ | 0.7463 | 0.033 | 22.698 | 0.000 | [0.682 0.811] |
| $x_3$ | 15.7827 | 1.937 | 8.149 | 0.000 | [11.974 19.592] |

The results of the multilinear regression (Table S. 2.) are shown in patients without rejection from the training set in Figure S. 2.  By using the same coefficients in patients with one rejection and multiple rejections, the regression lines are shown in Figure S. 3 and Figure S. 4.

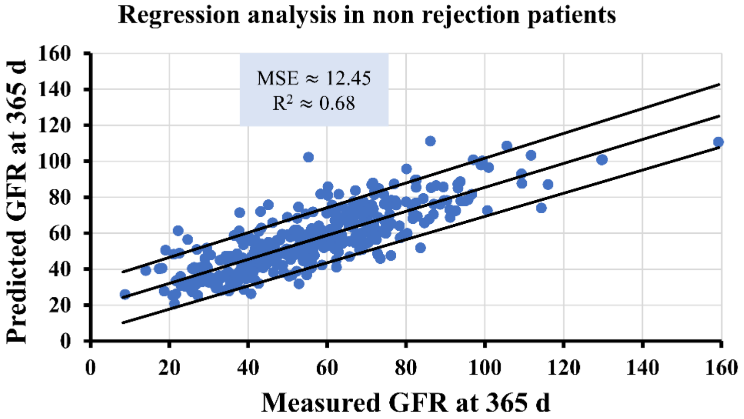


**Figure S. 2:**  Multi-linear regression with the predicting interval for the GFR at 365 d in patients without rejection (training set)

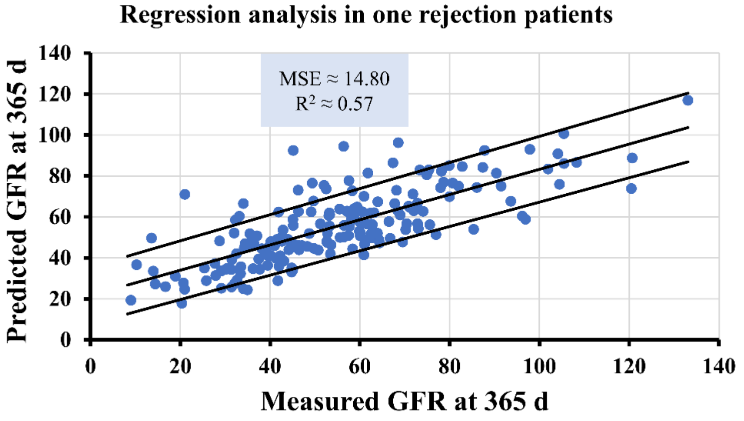


**Figure S. 3:**  Multi-linear regression with the predicting interval for the the GFR at 365 d in patients with one rejection (training set).

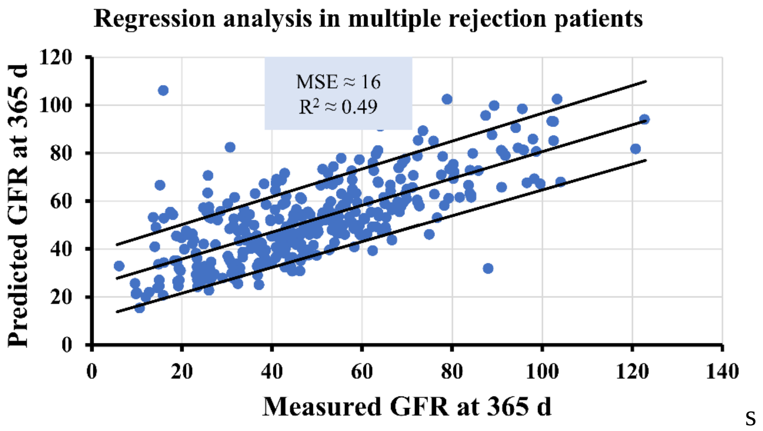


**Figure S. 4:** Multi-linear regression with the predicting interval for the the GFR at 365 d in patients with multiple rejections (training set).

The results showed that linear regression can be used for GFR prediction at 365 days posttransplantation in patients without rejection whereas both MSE and $R^2$ significantly decrease in patients with one and with multiple rejection.

### Section 3: Annual GFR decrease in real patients.

Based on Table S. 3 if patients with relevant complications which are known to affect the graft (rejection, BK virus nephropathy, glomerulonephritis, severe infections and other severe extra-renal diseases and urinary tract infections) are excluded the average annual loss is 1.58 ml/min 1.73 $m^2$/yr.

**Table S. 3:** Average loss per year in patients without rejection, BK virus nephropathy, glomerulonephritis, severe infections and other severe extra-renal diseases and urinary tract infections.

| | | Percentile | | | | | | |
|---|---|---|---|---|---|---|---|---|
| | | **5** | **10** | **25** | **50** | **75** | **90** | **95** |
| **Grounded method** | **Average loss per year** | -11.2680 | -7.4477 | -3.7441 | -1.5803 | 0.9553 | 3.4796 | 5.2999 |
| **Tukey test** | **Average loss per year** | | | -3.6809 | -1.5707 | 1.0690 | | |

Based on Table S. 4, the whole cohort of patients has an average annual GFR loss of 2.07 ml/min/1.73 $m^2$/yr).

**Table S. 4:** Average loss per year in the whole training cohort of patients

| | | Percentile | | | | | | |
|---|---|---|---|---|---|---|---|---|
| | | **5** | **10** | **25** | **50** | **75** | **90** | **95** |
| **Grounded method** | **Average loss per year** | -17.8305 | -10.6935 | -4.7898 | -2.0732 | 0.0798 | 1.9119 | 3.5672 |
| **Tukey test** | **Average loss per year** | | | -4.7898 | -2.0587 | .0798 | | |

From Eqs. S3 and S4 in the Section 1, it is assumed that $\frac{du}{dt} \approx 0$, $u$ = -2.07 ml/min/1.73 m$^2$/365 d, $\alpha$ =0, $b$ = 0. Also, if we considered $\lambda$ = 1 d$^{-1}$ and an average value of GFR equal to 60 ml/min/ 1.73 m$^2$, then substituting all these values in Eq. S.3,

$$\lambda u \approx -\gamma\, GFR_{ave} \Leftrightarrow \gamma = -\frac{\lambda u}{GFR_{ave}} \Leftrightarrow \gamma = -\frac{1\, d^{-1}\left(-\frac{2.07}{365}\,\frac{mL}{\min 1.73\, m^2\, d}\right)}{60\,\frac{mL}{\min 1.73\, m^2}} \Leftrightarrow$$

$$\Leftrightarrow \gamma = 9.45{\cdot}10^{-5} \approx 10^{-4}\, d^{-2}$$

**Section 4: Distribution of** $\boldsymbol{\lambda}$**.**

The distribution of $\lambda$ obtained from the parameter estimation of 362 patients without rejection is depicted in in Figure S. 5.

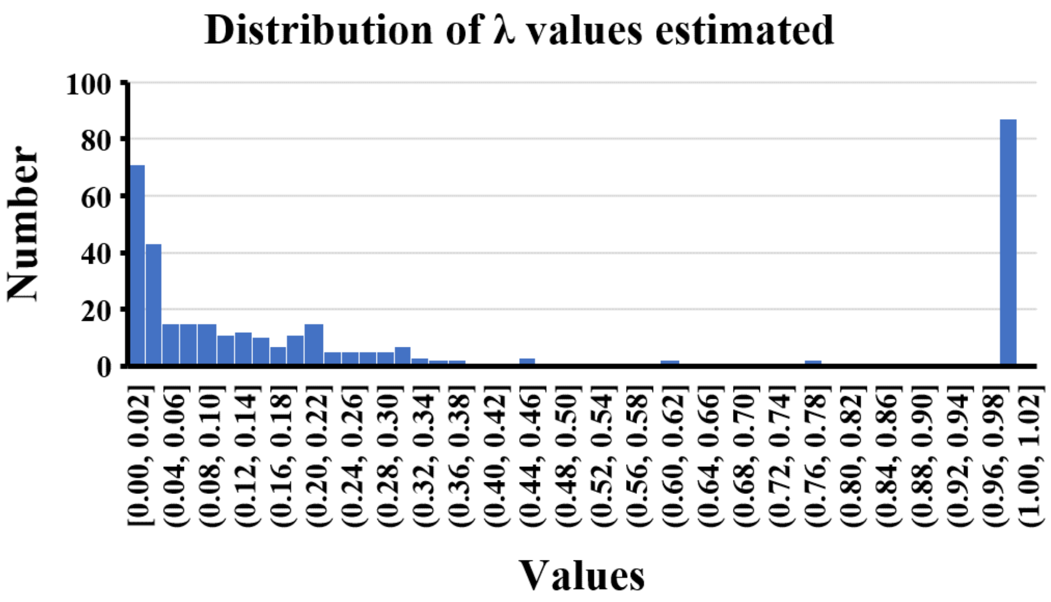


**Figure S. 5:** Distribution of λ values in the non-rejection patients.

Small values of $\lambda$ capture GFR courses that oscillate over time (underdamped oscillator)., whereas for high $\lambda$ the model could fit patients whose GFR course relaxes fast to a certain value (overdamped oscillator).

**Section 5: Graft Survival probabilities for all patients of the training group.**

The following data stem from an unpublished analysis which applied the same approach and cohort that was used in the study on patient survival from Scheffner et al (Scheffner et al., 2020)

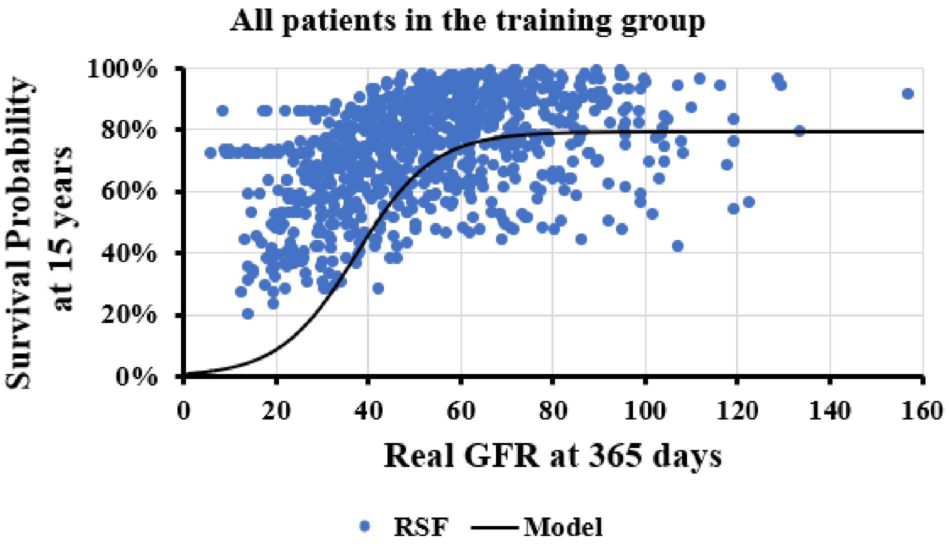


**Figure S. 6:** Model fitting of graft survival probability for all patients of the training group (Abbreviation: RSF is the random survival forest)

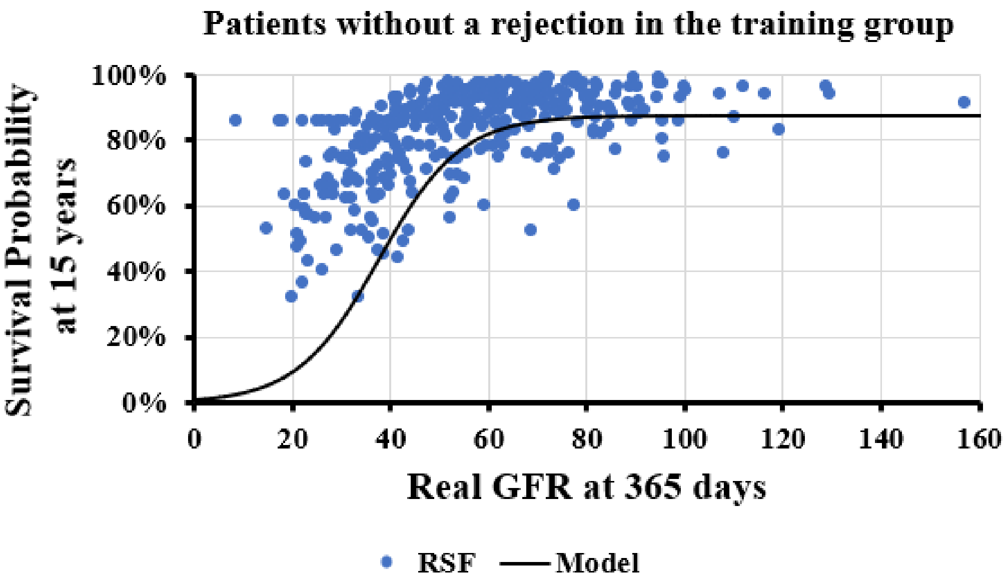


**Figure S. 7:** Model fitting of graft survival probability in patients without rejection in the training group (Abbreviation: RSF is the random survival forest)

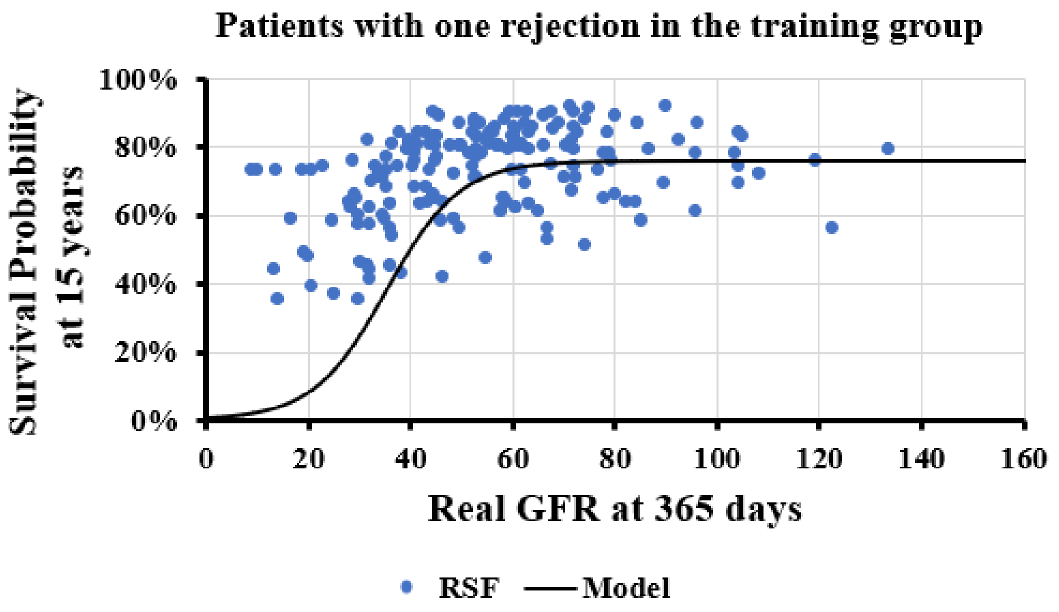


**Figure S. 8:** Model fitting of graft survival probability for patients with one rejection of the training group (Abbreviation: RSF is the random survival forest)

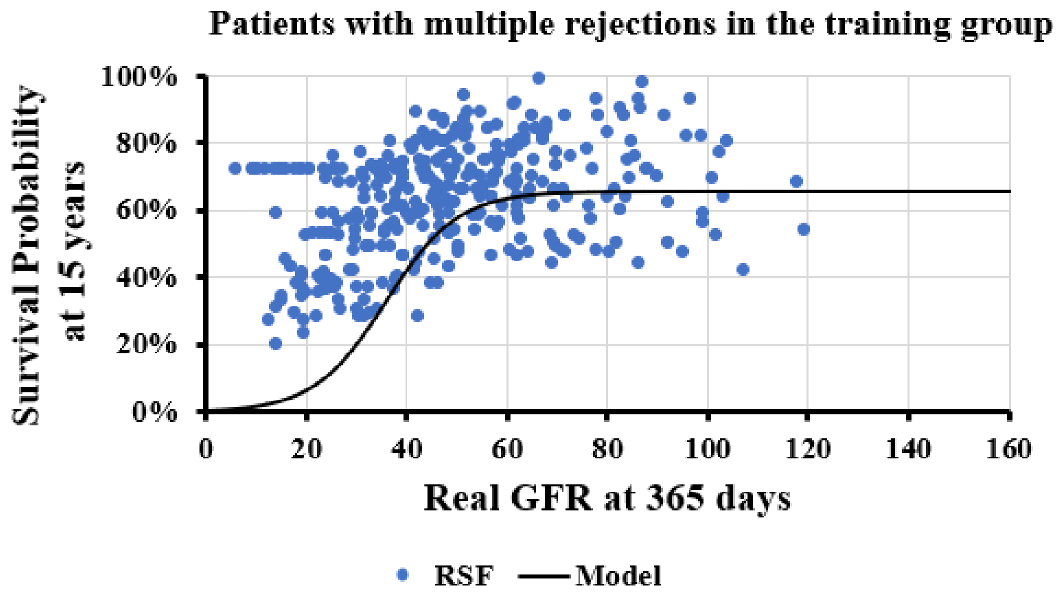


**Figure S. 9:** Model fitting of graft survival probability of patients with multiple rejections of the training group (Abbreviation: RSF is the random survival forest)

**Section 6: Classification with regular XGBoost, and Random Forest.**

In this section, the prediction of high risk patients was treated as a very simple classification problem. The high risk patients were those whose GFR annual value was less than a threshold The thresholds obtained from section 5 of SI were 30, 40, and 50 mL/min/1.73m$^2$.

$$failure = \begin{cases} 1, \ when\ measured\ GFR_{365\,d} \leq Clin_{thr} \\ 0, \ when\ measured\ GFR_{365\,d} > Clin_{thr} \end{cases} \quad \text{(S. 17)}$$

For classification XGBoost and Random Forest were performed in the patient with one, with multiple and without rejection, and the SMOTE (Synthetic Minority Over-sampling Technique) algorithm was used due to imbalanced data. The results are shown in Table S. *5*.

**Table S. 5:** XGBoost, and Random Forest accuracies for different critical thresholds ($Crit_{thr}$) in non-rejection patients.

| Classifier | AUC for $Crit_{thr}$ equal to 30 mL/min/1.73m$^2$ | AUC for $Crit_{thr}$ equal to 40 mL/min/1.73m$^2$ | AUC for $Crit_{thr}$ equal to 50 mL/min/1.73m$^2$ |
|---|---|---|---|
| **XGBoost** | 0.65 | 0.68 | 0.72 |
| **Random Forest** | 0.57 | 0.86 | 0.82 |

**Table S. 8:** XGBoost, and Random Forest accuracies for different critical thresholds ($Crit_{thr}$) in one rejection patients.

| Classifier | AUC for $Crit_{thr}$ equal to 30 mL/min/1.73m$^2$ | AUC for $Crit_{thr}$ equal to 40 mL/min/1.73m$^2$ | AUC for $Crit_{thr}$ equal to 50 mL/min/1.73m$^2$ |
|---|---|---|---|
| **XGBoost** | 0.72 | 0.77 | 0.80 |
| **Random Forest** | 0.73 | 0.73 | 0.75 |

**Table S. 9:** XGBoost, and Random Forest accuracies for different critical thresholds ($Crit_{thr}$) in multiple rejection patients.

| Classifier | AUC for $Crit_{thr}$ equal to 30 mL/min/1.73m$^2$ | AUC for $Crit_{thr}$ equal to 40 mL/min/1.73m$^2$ | AUC for $Crit_{thr}$ equal to 50 mL/min/1.73m$^2$ |
|---|---|---|---|
| **XGBoost** | 0.86 | 0.70 | 0.71 |
| **Random Forest** | 0.75 | 0.76 | 0.77 |

Based on the results on Table S. *5*-S.9, regardless of considering the issue of data imbalance, the classification accuracy depends on the threshold, and the accuracy is low especially when the threshold decreases.

### References

Scheffner, I., Gietzelt, M., Abeling, T., Marschollek, M., & Gwinner, W. (2020). Patient Survival after Kidney Transplantation: Important Role of Graft-sustaining Factors as Determined by Predictive Modeling Using Random Survival Forest Analysis. *Transplantation*, 1095–1107. https://doi.org/10.1097/TP.0000000000002922
